# Validation of the international consensus group criteria for slide review following automated complete blood count at Jimma Medical Center, Ethiopia

**DOI:** 10.1101/2022.10.26.22281576

**Authors:** Girum Tesfaye Kiya, Aklilu Getachew Mamo, Sintayehu Asaye Biya, Dejene Gebre Gobena, Natal Demeke, Tilahun Yemane Shenkutie

**Affiliations:** School of Medical Laboratory Science, Health Institute, Jimma University, Jimma, Ethiopia; Jimma Medical Center, Jimma University, Jimma, Ethiopia

## Abstract

**Background:** The international consensus group suggested criteria for action following automated complete blood count and white blood cell differential analysis. These criteria were set based on data from laboratories of developed countries. It is highly important to validate the criteria in developing countries where infectious diseases are still rampant that can affect blood cell count and morphology. Thus, this study was aimed to validate the consensus group criteria for slide review at Jimma Medical Center, Ethiopia from February 1 to May 30, 2020.

**Method:** The study comprised a total of 1685 patient samples from the daily laboratory workload of CBC analysis. The samples were collected in K2-EDTA tubes (Becton Dickinson) and analyzed using Coulter DxH 800 and Sysmex XT-1880 hematology analyzers. Slide review was done on two Wright-stained slides for each sample. All statistical analysis were performed using SPSS version 20 software.

**Result:** There were 39.8% positive findings, majority of which were related to red blood cells. The false negative and false positive rates for Sysmex and Coulter analyzer were 2.4% vs. 4.8%; and 4.6% vs. 4.7%, respectively. The false negative rate was unacceptably higher when we used physicians’ triggered slide review, which was 17.3% and 17.9% for Sysmex and Coulter analyzer, respectively.

**Conclusion:** Generally, the consensus group rules are suitable to use in our setting. However, we might still need to modify the rules, particularly to reduce the review rates. It is also necessary to confirm the rules with case mixes proportionally derived from the source population.

## Introduction

Automations in hematology laboratory changed the discipline dramatically as a result of analyzers which provide a reliable and accurate blood cell count, as a complete blood count (CBC) [1]. CBC is a test that measures the number and physical characteristics of cells in the circulation which are White blood cells (WBC), Red blood cells (RBC) and platelets, to determine general health status of the patient and screen, diagnose and monitor a variety of diseases and conditions that affect blood cells [2].

Generally, there are nine components of CBC named as white WBC count, RBC count, Hemoglobin (Hgb), hematocrit (Hct), mean corpuscular volume (MCV), mean corpuscular Hgb (MCH), mean corpuscular Hgb content (MCHC), platelet count, and red cell distribution width (RDW). Depending on the type of analyzer, some of these parameters are directly measured such as WBC, RBC, Hgb and MCV while others are calculated such as Hct [3]. Abnormalities in one or more components may indicate presence of one or more conditions which often leads to other tests to determine the cause of abnormalities, usually the Peripheral blood film (PBF) examination [2].

A PBF is a laboratory procedure that involves cytology of peripheral blood cells smeared on a slide. The diagnostic importance of PBF is enormous and its relevance has not been lessened by advances in hematology automations [4]. It is an inexpensive but powerful diagnostic tool and in some cases is sufficient to establish diagnosis alone [5].

However, some evidence recommended that there is a necessity of reducing PBF examination in light of time and monetary saving [6] and others suggested that the CBC and PBF are well correlated and hence usage of automation is recommended to ease workload and time for patients [7].

On the other hand, there are evidences that showed a significant discrepancy between automations and PBF on some parameters and suggested the interpretation of CBC results in light of the PBF examination [8], [9]. In addition to its importance in quality assurance to confirm findings of automations, PBF is found to be crucial to solely detect some important morphologic changes [10].

In Jimma Medical Center (JMC), PBF following CBC is performed without any objective criteria and mainly triggered by physicians. In 2005, the international consensus group (ICG) led by Dr. Berend Houwen suggested criteria for action following automated CBC and WBC differential analysis [11]. The group indicated the importance of these criteria for a wide range of Hematology laboratories and suggested validation of the rules in individual laboratories before implementation as one size does not fit all due to various reasons like practice of individual clinics, test volume, the instrument used and others. Moreover, the ICG criteria were established based on data from laboratories of developed countries including US, Canada, Australia, and Europe. Thus, it is highly important to validate the criteria in developing countries where infectious diseases are still rampant that can affect blood cell count and morphology. Thus, the current study was aimed to validate the consensus group criteria for slide review following automated CBC at JMC, Ethiopia.

## Materials and Methods

### Study setting

The study was conducted at JMC Hematology laboratory. The Medical Center is one of the largest health facilities in the country providing service to 15 million people with 1600 staff members and 800 beds. The average daily CBC analysis is about 200. The study was conducted from February 1 to May 30, 2020 comprising a total of 1685 blood samples from the daily laboratory workload of CBC analysis. The minimal sample size suggested by the international society for laboratory hematology (ISLH) was 1000 [11]. An average of 20 samples were randomly selected from the daily workload. The samples were collected in K2-EDTA tubes (Becton Dickinson) and analyzed using Coulter DxH 800 and Sysmex XT-1800 hematology analyzers. The samples were collected from more than 13 wards including Medical, outpatient, emergency, pediatrics, maternity, and oncology. About a fourth of all samples (27%) came from oncology department whilst samples from ICU and ART clinic were the least frequent.

From the total samples, 45% were from males and 55% were from females. The age of the patients ranges from 1 to 90 years. Among the total sample, 792 samples were analyzed by Sysmex XT-1800 while 893 were analyzed by Coulter DxH 800 hematology analyzers.

### Study Design

A cross-sectional study design was employed to validate the ICG criteria and to compare the criteria with the existing practice.

### Peripheral blood film review

Manual slide review was done on two Wright-stained slides for each sample by two experienced technicians, independently and blinded to the automation results. Positive results were defined based on international society for laboratory hematology (ISLH) recommendation (Table 1).

**Table 1:**
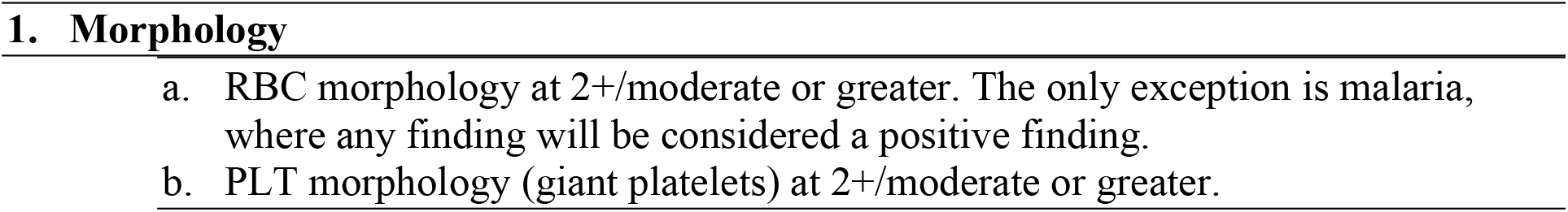
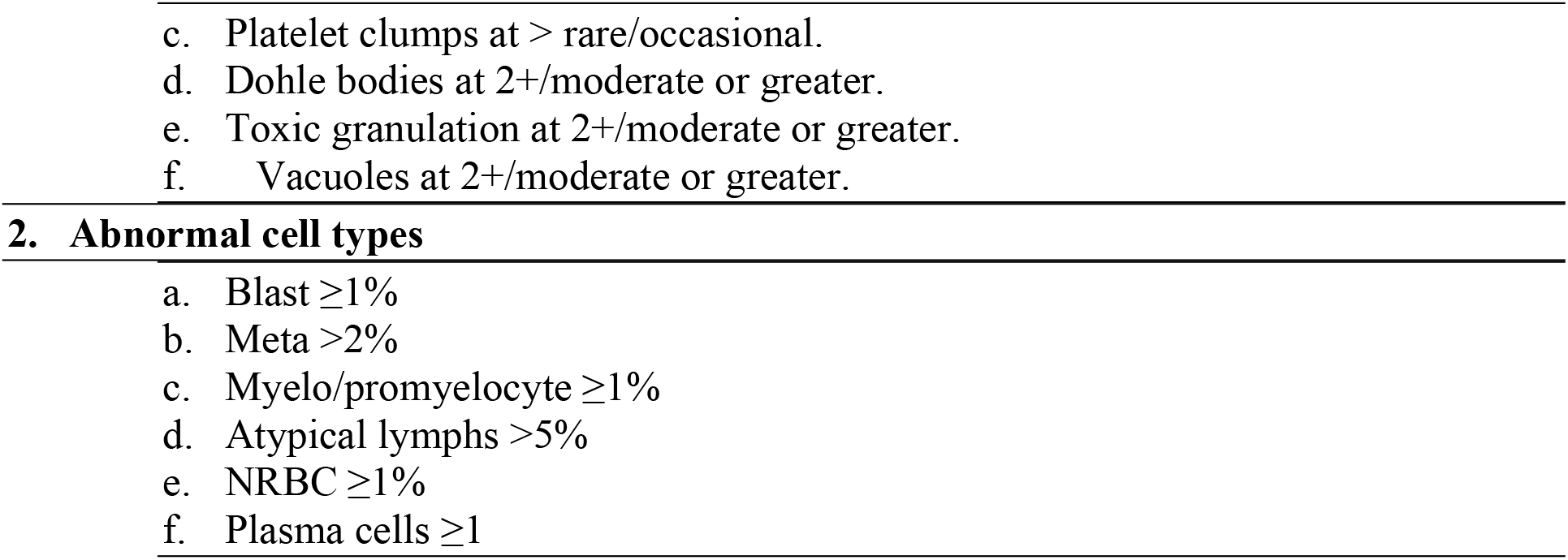
Criteria for a positive smear [11]

### Statistical analysis

All statistical analysis were performed using SPSS version 20 software. The automated CBC results were sorted in to four categories as a truth table based on correlation between the positive/ negative status defined by the ICG rules versus the “truth” defined by the slide review. If a rule was triggered, and the smear contained a positive finding, the sample was graded as true positive (TP). If a rule was triggered, but the smear did not contain any positive findings, the sample was graded as false positive (FP). True negative (TN) grade was given when there was no triggered rule, and the smear did not contain any positive finding. False negative (FN) grade was given when there was no triggered rule, but the smear contained a positive finding [11].

Sensitivity, specificity, accuracy rate, error rate, review rate, positive predictive values and negative predictive values were calculated based on the following calculation:

|  |  |
| --- | --- |
| Sensitivity = | $TP/(TP+FN)$ |
| Specificity = | $TN/(FP+TN)$ |
| Accuracy rate = | $(TP+TN)/Total$ |
| Error rate = | $(FP+FN)/Total$ |
| Review Rate = | $(TP+FP)/Total$ |
| Predictive Value of a Positive Result = | $TP/(TP+FP)$ |
| Predictive Value of a Negative Result = | $TN/(FN+TN)$ |

### Ethical considerations

Ethical clearance was obtained from Jimma University, Institute of Health Ethical Review Committee. Support letter from Health science research coordinating office was written to JMC. Permission was obtained from medical director office of the medical center. Any information concerning the participants was kept confidential and the analysis done for the participants was used only for the intended purposes. Authors had no access to information that could identify individual participants during or after data collection. Patient result of the slide review was communicated to the treating physician.

## Result

### Analysis of Smear review findings

Out of the total 1685 samples, there were 671 (39.8%) positive findings according to the definition of ISLH; 283 (42%) were positive samples from Coulter analyzer whereas 388 (58%) were positive samples from Sysmex analyzer. Majority of the abnormalities were related to RBC, followed by WBC and platelet abnormalities. Anisocytosis was the most common finding while giant platelet is the least common.

### Analysis of the international consensus group criteria

The truth table showed that FN and false positive FP results were 2.4% and 4.8%, respectively for Sysmex analyzer. For Coulter analyzer, the FN and FP rates were 4.6% and 4.7%, respectively. The FN rate was within the acceptable range for both analyzers which is less than 5%. The review rate was also found to be 51% and 31.8% for the two analyzers, respectively. The diagnostic performance of both analyzers based on the ICG criteria and slide review positivity was found to be good with sensitivity and specificity of 95.1% and 90.6% on Sysmex and 85.5% and 93.1% on Coulter analyzer (Table 2).

**Table 2:** Truth table based on the international consensus group criteria

| Status | Instruments |  |
| --- | --- | --- |
|  | Sysmex | Coulter |
| True negative | 365 (46.1%) | 564 (63.2%) |
| True positive | 369 (46.6%) | 242 (27.1%) |
| False negative | 19 (2.4%) | 41 (4.6%) |
| False positive | 38 (4.8%) | 42 (4.7%) |
| Review rate | 407 (51%) | 284 (31.8%) |
| Sensitivity | 95.1% | 85.5% |
| Specificity | 90.6% | 93.1% |
| Positive predictive value | 90.6% | 85.2% |
| Negative predictive value | 95.1% | 93.2% |
| Accuracy rate | 92.7% | 90.3% |
| Error rate | 7.2% | 9.3% |

### False Negative

The total number of FN occurrences on Sysmex and Coulter analyzer were 24 and 57 and the total number of FN cases were 19 and 41, respectively. Majority of the occurrences on samples from coulter analyzer and Sysmex analyzer were related to RBC abnormalities, particularly, anisocytosis followed by findings of macrocytes. Moreover, white blood cell abnormalities such as toxic granulation and atypical lymphocyte contributed to the FN rate (Fig. 1).

**Figure 1.**
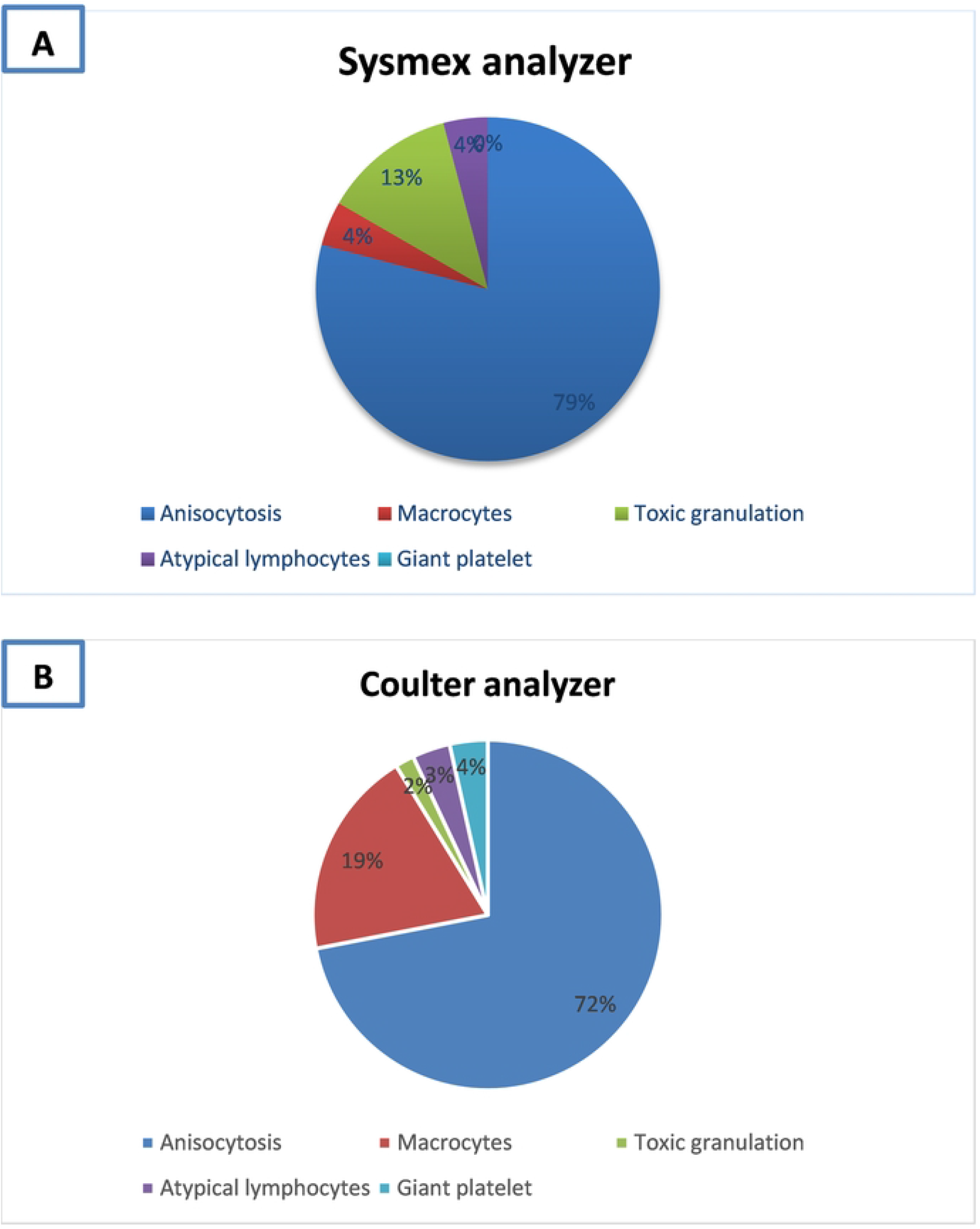

### False positive and True positive

The most frequently triggered rule on Sysmex analyzer was rule number 5 followed by rule no. 7, 9, and 10. Rule no. 7 was the most frequently triggered rule on Coulter analyzer followed by rule no. 9, 10, and 5. Generally, the TP rate was higher on Sysmex analyzer than Coulter analyzer. In total, 18 cases triggered rule no. 5 (WBC count) on Sysmex analyzer, majority (17/18) of which appears to be because of the neutrophil count (rule no. 17). The same is true on Coulter analyzer where 10 cases triggered rule no. 5 and 19 cases triggered rule no. 17. The second most common triggered rule among the FP cases was rule no. 7 (platelet count) which was 17 and 16 cases on Sysmex and Coulter analyzer, respectively (Table 3).

**Table 3.** True positive and False positive

| Rule no. | Parameters | Rules | Sysmex |  | Coulter |  |
| --- | --- | --- | --- | --- | --- | --- |
|  |  |  | FP rate, n (%) | TP rate, n (%) | FP rate, n (%) | TP rate, n (%) |
| 5 | WBC | $<4 \times 10^9/L$ or $>30 \times 10^9/L$ | 18 (47.4) | 141 (38.4) | 10 (23.8) | 80 (33) |
| 7 | PLT | $<100 \times 10^6/L$ or $>1000 \times 10^6/L$ | 17 | 140 | 16 | 105 |
| 9 | HGB | $<7g/dl$ or $>2g/dl$ from upper reference range for age and sex | 9 | 135 | 3 | 92 |
| 10 | MCV | $<75fl$ or $>105fl$ | 6 | 130 | 8 | 88 |
| 17 | Neutrophil | $<2 \times 10^9/L$ or $>20 \times 10^9/L$ | 17 | 121 | 19 | 77 |
| 15 | RDW | $>22$ | 8 | 87 | 3 | 65 |

WBC: White blood cell; PLT: Platelet; HGB: Hemoglobin; MCV: Mean cell volume; RDW: Red cell distribution width; FP: False positive; TP: True positive

### Comparison between the consensus group criteria and physicians’ triggered slide review

From the total sample included in the study, 616 (36.5%) slide reviews were triggered by physicians. The truth table based on physicians’ triggered slide review showed different result on both analyzers. The FN rate was unacceptably higher on both analyzers as compared to the ICG criteria (Fig. 2). The review rate was also found to be 49% and 25.5% on Sysmex and Coulter analyzers, respectively.

**Figure 2.**
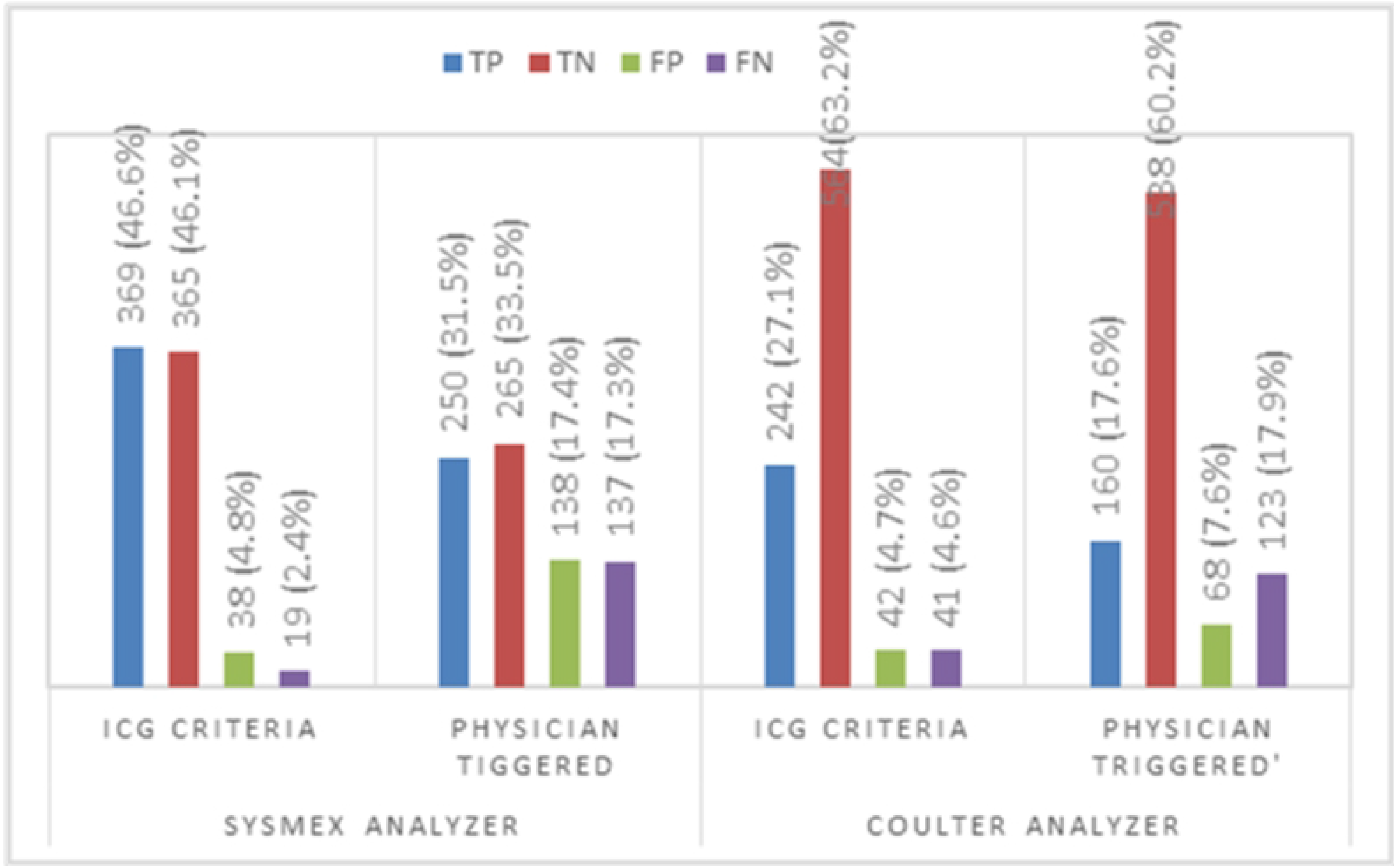

## Discussion

A consensus group established by the ISLH suggested indications for slide review and/or manual WBC differential count following automated CBC [11]. The group was comprised of 20 experts from multiple developed countries who use multiple instruments. Criteria at participants’ laboratories were discussed and finally combined in to 41 rules. These rules have to be tested in different laboratories in the world before use. The present study involved 1685 samples analyzed by two automated instruments at JMC in Ethiopia to test if the recommended criteria are well fitting to laboratories of developing countries.

In this study, evaluation of the ICG review criteria yielded a FN rate of 2.4% and 4.6% for Sysmex and coulter analyzers, respectively. These results were within the acceptable range according to the ICG recommendation of a maximum acceptable FN rate less than 5% [11]. Though anisocytosis is the most common finding that caused FN result, adjustment of rule #15 (RDW rule) may not be necessary as the rate is within the acceptable range. The FP rate on both analyzers was also acceptable when compared to the rate that was found by the ICG, which is 18.6%. Rule #5 and # 17 were the most frequent triggered rules that caused FP result. Increased WBC count (>30 ×10^9^/L) rather than decreased WBC (<4 ×10^9^/L), particularly the neutrophil count (>20 ×10^9^/L) was the frequent reason as expected in the areas where bacterial infection is common [12].

The college of American Pathologists (CAP) had Q-probe study of 95,151 CBC determinations performed in 263 institutions and the median PBF review rate was 26.7%, which was suggested as a normative review rate [13]. The present study showed that the review rate for Sysmex and Coulter analyzer were 51% and 31.8%, respectively. The review rate of samples from Sysmex analyzer appear to be higher as compared to the review rate of the Q-probe study by CAP. However, a lower review rate, though desirable, might say nothing about quality. In our case, the TP rate was higher in samples from Sysmex analyzer as compared to the samples analyzed by Coulter. This might be due to the patient mix analyzed by this analyzer, majority of which are from oncology and emergency department, as compared to the samples analyzed by Coulter machine.

The comparison of our finding with previous published studies showed that the FN rate is higher than the previous findings, in which the rate range from 1.6 to 2.2 [14]-[15], but lower than studies which reported FN rate ranging from 7.37 to 9.25 [16], 19). The FP rate in this study is much better than previous studies which reported FP rate ranging from 7.2% to 34% [14], [18], [15], [17]. Moreover, the review rate in the present study was found to be comparable with some previous studies which reported a review rate ranging from 29% to 51% [14], [15],[17], [19].

This study has also compared the ICG criteria and physicians’ triggered slide review. In fact, the physicians’ triggered slide review practice appears to be arbitrary in a sense that no specific rule is set following automated CBC to indicate slide review. The truth table showed that FN rate was unacceptably higher among slide reviews triggered by physicians. This indicates that much positive findings are remaining unnoticed due to lack of objective rules which in turn might affect patients’ safety.

In conclusion, generally the ICG rules set by ISLH is suitable to use in our setting. However, we might still need to modify the rules, particularly to reduce the review rates as there is shortage of skilled technicians to review large amount of sample. On the other hand, though the review rate was desirably lower for samples from Sysmex analyzer, continuing with physicians’ triggered slide review approach might result in increased unnoticed positive findings. It is also necessary to confirm the rules with case mixes proportionally derived from the source population.

## Data Availability

All relevant data are within the manuscript and its Supporting Information files.

## Reference

[1] K. W. Jones, “Evaluation of Cell Morphology and Introduction to Platelet and White Blood Cell Morphology,” Philadelphia PA: FA. Davis. Company, 2010.

[2] “Complete blood count. Lab Tests Online,” 2022. https://labtestsonline.org.uk/tests/full-blood-count-fbc (accessed Jul. 07, 2022).

[3] C. Betty, Hematology in practice. F.A Davis Company, 2010.

[4] N. B. Adewoyin AS, “Peripheral blood film - a review,” Ann Ib Postgr. Med, vol. 12, no. 2, pp. 71–79, 2014.

[5] B. J. Bain, “Diagnosis from the blood smear,” N Engl J Med, vol. 353, p. 498, 2005.

[6] R. J. Joubert J, Weyers R, “Reducing unnecessary blood smear examinations: can Sysmex blood cell analysers help,” Med. Technol. SA, vol. 28, pp. 6–12, 2014.

[7] S. O. Ike, T. Nubila, E. O. Ukaejiofo, I. N. Nubila, E. N. Shu, and I. Ezema, “Comparison of haematological parameters determined by the Sysmex KX - 2IN automated haematology analyzer and the manual counts,” BMC Clin. Pathol., vol. 10, no. 1, pp. 1–5, Apr. 2010, doi: 10.1186/1472-6890-10-3/TABLES/2.

[8] N. M. Poonam R, “Automated red blood cell analysis compared with routine RBC morphology by smear review,” Blood, vol. 56, pp. 34–39, 2011.

[9] M. T. Aseem Jain, A. Bhake, “Co-Relative Study on Peripheral Blood Smears in Anemia with Automated Cell Counter Generated Red Cell Parameters,” Semant. Sch., 2018.

[10] D. S. Rosenthal, “Evaluation of the peripheral blood smear - UpToDate,” 2022. https://www.uptodate.com/contents/evaluation-of-the-peripheral-blood-smear?search=peripheralbloodfilm&usage_type=default&source=search_result&selectedTitle=1∼150&display_rank=1 (accessed Jul. 07, 2022).

[11] P. W. Barnes, S. L. McFadden, S. J. Machin, and E. Simson, “The international consensus group for hematology review: suggested criteria for action following automated CBC and WBC differential analysis,” Lab. Hematol., vol. 11, no. 2, pp. 83–90, 2005, doi: 10.1532/LH96.05019.

[12] S. M. Honda T, Uehara T, Matsumoto G, Arai S, “Neutrophil left shift and white blood cell count as markers of bacterial infection,” Clin Chim Acta, vol. 457, pp. 46–53, 2016.

[13] D. A. Novis, M. Walsh, D. Wilkinson, M. St. Louis, and J. Ben-Ezra, “Laboratory productivity and the rate of manual peripheral blood smear review: a College of American Pathologists Q-Probes study of 95,141 complete blood count determinations performed in 263 institutions,” Arch. Pathol. Lab. Med., vol. 130, no. 5, pp. 596–601, May 2006, doi: 10.5858/2006-130-596-LPATRO.

[14] B. Pratumvinit, P. Wongkrajang, K. Reesukumal, C. Klinbua, and P. Niamjoy, “Validation and optimization of criteria for manual smear review following automated blood cell analysis in a large university hospital,” Arch. Pathol. Lab. Med., vol. 137, no. 3, pp. 408–414, Mar. 2013, doi: 10.5858/ARPA.2011-0535-OA.

[15] S. Pipitone et al., “Comparing the performance of three panels rules of blood smear review criteria on an Italian multicenter evaluation,” Int. J. Lab. Hematol., vol. 39, no. 6, pp. 645–652, Dec. 2017, doi: 10.1111/IJLH.12720.

[16] A. S. Eldanasoury, N. H. Boshnak, R. E. Abd, and E. Monem, “Validation of Criteria for Smear Review Following Automated Blood Cell Analysis in Ain Shams University Laboratory,” Int. J. Sci. Res., vol. 5, pp. 2319–7064, 2013.

[17] S. R. Comar, M. Malvezzi, and R. Pasquini, “Are the review criteria for automated complete blood counts of the International Society of Laboratory Hematology suitable for all hematology laboratories?,” Rev. Bras. Hematol. Hemoter., vol. 36, no. 3, p. 219, Sep. 2014, doi: 10.1016/J.BJHH.2014.03.011.

[18] S. Buoro et al., “Validation rules for blood smear revision after automated hematological testing using Mindray CAL-8000,” J. Clin. Lab. Anal., vol. 31, no. 4, Jul. 2017, doi: 10.1002/JCLA.22067.

[19] X. Wang et al., “Establishment of improved review criteria for hematology analyzers in cancer hospitals,” J. Clin. Lab. Anal., vol. 35, no. 2, Feb. 2021, doi: 10.1002/JCLA.23638.

